# Relations between retinal microvasculature by optical coherence tomography angiography and cerebral small vessel disease in individuals with type 1 diabetes

**DOI:** 10.1101/2025.10.20.25338353

**Authors:** Aleksi Tarkkonen, Iiris Kyläheiko, Marika I Eriksson, Per-Henrik Groop, Lena M Thorn, Joni A. Turunen, Jukka Putaala, Juha Martola, Daniel Gordin, the FinnDiane Study Group

**Affiliations:** Department of Radiology, University of Helsinki and Helsinki University Hospital, Helsinki, Finland; Minerva Foundation Institute for Medical Research, Helsinki, Finland; Department of Psychology and Logopedics, Faculty of Medicine, University of Helsinki, Helsinki, Finland; Department of Neurology, Kymenlaakso Central Hospital, Kotka, Finland; Folkhälsan Research Center, Helsinki, Finland; Department of Nephrology, University of Helsinki and Helsinki University Hospital, Helsinki, Finland; Research Program for Clinical and Molecular Metabolism, Faculty of Medicine, University of Helsinki, Helsinki, Finland; Department of General Practice and Primary Health Care, University of Helsinki and Helsinki University Hospital, Helsinki, Finland; Department of Ophthalmology, University of Helsinki and Helsinki University Hospital, Helsinki, Finland; Eye Genetics Group, Folkhälsan Research Center, Biomedicum Helsinki, Helsinki, Finland; Neurology, Helsinki University Hospital and University of Helsinki, Helsinki, Finland; Joslin Diabetes Center, Harvard Medical School, Boston, MA, USA

**Keywords:** Type 1 Diabetes, Cerebral Small Vessel Disease, Optical Coherence Tomography Angiography

## Abstract

**Backround:** Nearly all individuals with type 1 diabetes develop diabetic retinopathy over time, known to relate to cerebral small vessel disease. We investigated associations between early vascular changes in the eye and brain by optical coherence tomography angiography (OCTA) and brain magnetic resonance imaging (MRI) in middle-aged, neurologically asymptomatic individuals with type 1 diabetes.

**Research Design and Methods:** Individuals with type 1 diabetes (n=159, median age 48.0 years, diabetes duration 29.3 years, 53% female) and 49 healthy controls underwent clinical and biochemical assessments, brain MRI to evaluate cerebral microbleeds (CMBs) and white matter hyperintensities (WMHs), and macular 3×3 mm OCTA imaging to assess vessel densities of superficial (SCP) and deep (DCP) retinal capillary plexuses, and the area of the foveal avascular zone (FAZ).

**Results:** No differences in OCTA parameters were observed between individuals with type 1 diabetes and healthy controls. However, SCP and DCP vessel densities (45.1% vs 46.5%, p=0.034, and 49.9% vs 52.9%, p<0.001, respectively), were smaller in individuals with type 1 diabetes with CMBs compared with those without. No such evidence was found for FAZ. In univariate linear regression models, SCP and DCP vessel densities were negatively associated with age, diabetes duration, blood pressure, kidney function (eGFR), number of CMBs, and WMHs. FAZ was related to diabetes duration, age of diabetes onset, and LDL cholesterol concentration. Of these, in multivariate models, diabetes duration remained associated negatively with vessel densities and positively with FAZ (SCP: standardized β =-0.210 [p=0.014]; DCP: standardized β =-0.275 [p<0.001]; FAZ: standardized β=0.295 [p<0.001]). None of the retinal markers differed in individuals with or without WMHs and type 1 diabetes.

**Conclusion:** In middle-aged, neurologically asymptomatic adults with long-standing type 1 diabetes, lower SCP and DCP vessel densities were associated with cerebral microbleeds, suggesting OCTA may aid cerebrovascular risk stratification but not WMH assessment.

**Research insights:** *What is currently known about this topic?:* Nearly all individuals with type 1 diabetes develop diabetic eye disease within 30 years following the diagnosis. We observed previously that one third of neurologically asymptomatic individuals with type 1 diabetes have cerebral microbleeds in routine brain magnetic resonance imaging. Cerebral microbleeds were more common among those with advanced forms of diabetic eye disease.

*What is the key research question?:* To investigate associations between markers of cerebral small vessel disease and metrics of optical coherence tomography angiography among individuals with type 1 diabetes.

*What is new?:* Vessel densities of the macula were smaller among those individuals with type 1 diabetes and cerebral microbleeds. Vessel densities were negatively and area of the foveal avascular zone positively associated with duration of diabetes.

*How might this study influence clinical practice?:* OCTA may aid cerebrovascular risk stratification among individuals with long-standing type 1 diabetes

## Backround

Individuals with type 1 diabetes have an increased risk for microvascular complications, such as diabetic retinopathy (DR) and cerebral small vessel disease (cSVD). In neurologically asymptomatic, middle-aged individuals with type 1 diabetes, we have earlier showed particularly white matter hyperintensities (WMHs) and cerebral microbleeds (CMBs) in brain MRI ^1,2^.

The brain and retina originate from the ectoderm and are hence closely connected anatomically and physiologically and share vascular origins ^3–5^. Indeed, we observed earlier that even one-third of middle-aged neurologically asymptomatic individuals with type 1 diabetes had CMBs ^1,2^, and that CMBs were more common among those individuals with severe nonproliferative or proliferative DR ^6^.

Optical coherence tomography angiography (OCTA) enables noninvasive, objective, layer-specific analysis of the retinal microvasculature, including the most commonly studied markers of superficial capillary plexus (SCP) and deep capillary plexus (DCP), as well as the area of the foveal avascular zone (FAZ) ^7,8^. Studies have reported differences in these OCTA parameters among individuals with type 1 diabetes compared to healthy controls already before clinical signs of diabetic retinopathy ^9,10^, as well as in diabetic retinopathy ^11,12^.

In studies conducted in older individuals with sporadic cSVD and not focusing specifically on individuals with type 1 diabetes, WMHs have been associated with OCTA markers^13–16^, but CMBs have not ^13,16,17^. However, no evidence exists on whether retinal OCTA markers, representing an extension of the central nervous system, and the cSVD brain markers are linked with each other in middle-aged individuals with type 1 diabetes.

We aimed to assess whether retinal microvascular OCTA markers and cSVD brain findings are associated in middle-aged adults with type 1 diabetes who have no overt neurological symptoms. Additionally, we compared retinal microvascular pathology in individuals with type 1 diabetes and healthy controls.

## Methods

This study is part of the prospective Finnish Diabetic Nephropathy (FinnDiane) Study, described in detail previously ^18^. The study follows the Declaration of Helsinki, with approval from the Ethics Committee of Helsinki University Hospital. Written informed consent was obtained from all participants. Between 2011 and 2017, 191 individuals with type 1 diabetes were enrolled in this sub-study investigating covert cerebrovascular disease in neurologically asymptomatic individuals with type 1 diabetes. Eligible participants were between 18 and 50 years of age, they had no history or symptoms of neurological disease, no kidney replacement therapy, nor any contraindications to MRI ^1^. Of the remaining 187 participants, 172 (92%) attended the follow-up visit in 2019 to 2024. There were no differences in clinical characteristics between the participants who attended the follow-up visits and those who did not (data not shown). Five individuals were excluded (multiple sclerosis, n=2, diffuse axonal injury, n=3). OCTA scans of sufficient quality were obtained for 159 individuals. Additionally, 49 healthy control participants matched by age, sex, and education, were enrolled at the follow-up phase.

## Clinical assessments

Clinical and laboratory assessments were done at the Helsinki FinnDiane research unit. This encompassed anthropometrics, blood pressure (an average of two blood pressure measurements was used), medical history, and lifestyle queries. Plasma creatinine, lipids, lipoproteins, HbA_1c_, and high-sensitivity C-reactive protein (hs-CRP) were measured using standard biochemical methods. Albuminuria was defined as an increased albumin excretion rate (≥20 μg/min or ≥30 mg/24h) in two of three urine collections. The Chronic Kidney Disease Epidemiology Collaboration formula was used to estimate glomerular filtration rate^19^.

## Optical coherence tomography angiography (OCTA)

OCTA images were captured using the SOLIX 1.0 (Optovue Inc., Fremont, CA, USA), ensuring a signal strength of 55 or higher, with a lateral resolution of 15 μm and an axial resolution of 5 μm, employing the AngioVue® 3 x 3 mm protocol. Both eyes were scanned within a 3×3 mm area centered on the fovea. Vessel density measurements for SCP and DCP, as well as FAZ, were performed using automated software with Projection Artifact Removal (3D PAR). Macular edema was evaluated to control it as a potentially confounding factor in the analysis visually by an experienced ophthalmologist (J.A.T). The right eye was used for analysis unless image quality was <7, in which case the left eye was used. Participants with insufficient image quality of both eyes were excluded.

## Brain MRI

Brain MRI scans were conducted using 3T Achieva (Philips Ingenia, the Netherlands) at the Medical Imaging Center of Helsinki University Hospital ^2^, within one year of the participants’ clinical and OCTA evaluations. The imaging protocol included multiple sequences: T1-weighted, T2-weighted, fluid-attenuated inversion recovery (FLAIR), susceptibility-weighted imaging, T2*, diffusion-weighted imaging (DWI), T1 magnetization-prepared rapid gradient echoand magnetic resonance time-of-flight. A senior neuroradiologist (J.M.), blinded to all clinical, OCTA, and biochemical data, assessed the scans for CMBs, WMHs, and infarcts, using established standardized criteria ^20,21^.

## Statistical Analysis

Continuous variables were assessed using the Wilcoxon test. Categorical variables were analyzed using the χ2 test. Results are reported as medians with interquartile ranges for descriptive statistics. Univariate linear regression analyses were applied to explore associations between OCTA parameters and clinical and selected brain MRI markers. Based on statistically significant univariate associations, multivariate linear regression models were developed to investigate these associations further. Model assumptions were assessed using standard diagnostic plots and statistical tests for normality, homoscedasticity, and multicollinearity. Age of onset was excluded from the analyses, as it represents only the difference between age and diabetes duration. As sensitivity analyses, statistical analyses were also conducted excluding participants with macular edema. Missing values were not replaced. Results are reported as standardized betas. Statistical significance was defined as p<0.05. Analyses were conducted using R (version 4.1.1).

## Results

### Clinical characteristics

Clinical characteristics of individuals with type 1 diabetes and healthy controls are presented in Table 1. Of those with type 1 diabetes, 51 (31.4%) had a Fazekas grade 0, 106 (66.7%) had Fazekas grade 1, and 2 (1.3%) had Fazekas grade 2. Out of controls, 11 (22.4%) had Fazekas grade 0, 36 (73.5%) had Fazekas grade 1, and 1 Fazekas grade (2.0%). Macular oedema was observed in 13.8% (22/159) of individuals with type 1 diabetes, and none of the controls. The median difference between MRI and OCTA scanning was 18 days, with an interquartile range of 5 to 44 days.

**Table 1.** Clinical characteristics.

| Variable | Participants with type 1 diabetes | Controls | P |
| --- | --- | --- | --- |
| n | 159 | 49 |  |
| Female sex, % | 84 (53) | 25 (52) | 0.617 |
| Age, years | 48.0 (41.5-53.4) | 49.3 (40.4-54.9) | 0.635 |
| Diabetes duration, years | 29.3 (25.7-37.8) | - | - |
| Age at onset, years | 13.9 (7.8-23.5) | - | - |
| Body mass index, kg/m<sup>2</sup> | 27.3 (24.9-30.4) | 25.4 (23.5-27.0) | 0.002 |
| Systolic blood pressure, mmHg | 130 (118-141) | 126 (119-137) | 0.373 |
| Diastolic blood pressure, mmHg | 82 (75-87) | 82 (78-89) | 0.195 |
| HbA1c, mmol/mol | 59 (51-66) | 34 (32-36) | <0.001 |
| Creatine, µmol/l | 66 (57-76) | 69 (60-79) | 0.174 |
| Total cholesterol, mmol/l | 4.0 (3.4-4.6) | 4.80 (4.3-5.4) | <0.001 |
| HDL Cholesterol, mmol/l | 1.42 (1.12-1.72) | 1.37 (1.15-1.61) | 0.462 |
| LDL Cholesterol, mmol/l | 2.07 (1.65-2.82) | 2.99 (2.56-3.45) | <0.001 |
| Triglycerides, mmol/l | 0.87 (0.68-1.26) | 0.93 (0.76-1.13) | 0.409 |
| eGFR, ml/min/1.73m<sup>2</sup> | 105 (95-111) | 101 (94-108) | 0.270 |
| hs-CRP (mg/l) | 1.1 (0.5-2.3) | 0.7 (0.4-2.2) | 0.061 |
| Retinal photocoagulation, n (%) | 51 (29) | 0 (0) | <0.001 |
| Antihypertensive medication, n (%) | 75 (47) | 6 (13) | <0.001 |
| Lipid-lowering medication, n (%) | 92 (58) | 2 (4) | <0.001 |
| Current smoker, n (%) | 7 (4.6) | 8 (16) | 0.770 |
| **MRI findings** |  |  |  |
| Cerebral SVD, n (%) | 111 (70) | 27 (56) | 0.116 |
| CMBs, n (%) | 56 (35) | 5 (10) | 0.002 |
| WMHs, n (%) | 98 (62) | 27 (56) | 0.617 |
| **OCTA findings** |  |  |  |
| Superficial vessel density, % | 45.99 (42.86-48.18) | 46.18 (43.97-47.04) | 0.983 |
| Deep vessel density, % | 52.20 (48.80-53.99) | 52.37 (49.81-53.59) | 0.718 |
| Foveal avascular zone, mm<sup>2</sup> | 0.28 (0.22-0.37) | 0.27 (0.21-0.33) | 0.433 |

Clinical characteristics, and brain magnetic resonance imaging (MRI) and optical coherence tomography angiography (OCTA) findings in participants with type 1 diabetes and controls.

Data are presented as n (%) or median (interquartile range). CMB, cerebral microbleed; eGFR, estimated glomerular filtration rate; HDL, high-density lipoprotein; hs-CRP, high-sensitivity C-reactive protein; LDL, low-density lipoprotein; SVD, small vessel disease; WMH, white matter hyperintensity.

### Associations between OCTA, clinical, and brain MRI parameters among individuals with type 1 diabetes

In univariate linear regression analyses (Table 2), both SCP and DCP vessel densities were associated negatively with age, diabetes duration, diastolic blood pressure, kidney function (eGFR), and WMHs. Both vessel densities associated positively with age of onset. DCP vessel density was additionally associated negatively with the number of CMBs. FAZ was associated positively with diabetes duration and serum LDL cholesterol concentration and negatively with age of onset. Only duration of diabetes remained significantly associated with SCP and DCP vessel densities and FAZ after adjusting for aforementioned variables, in multivariate analyses (standardized betas and p-values for SCP vessel density: r=-0.238 (p=0.006), DCP vessel density: r=-0.268 (p=0.003), and FAZ: r=0.219 (p=0.006)). These three associations remained even after excluding individuals with macular edema from the analyses (standardized betas and p-values for SCP vessel density: r=-0.289 (p=0.002), DCP vessel density: r=-0.244 (p=0.009), and FAZ: r=0.230 (p=0.008).

**Table 2.**
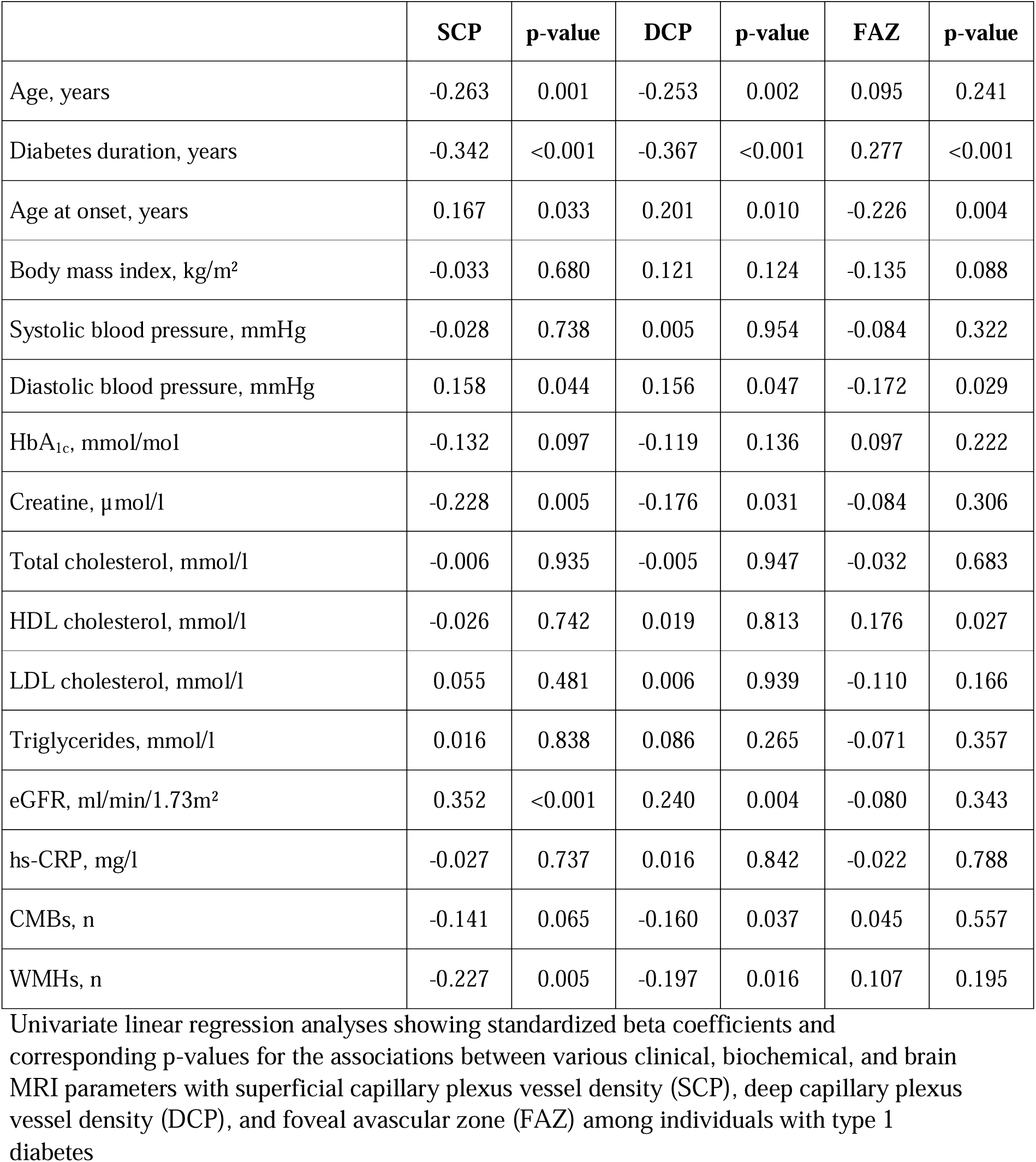
Standardized beta coefficients in univariate linear regression analyses.

|  | SCP | p-value | DCP | p-value | FAZ | p-value |
| --- | --- | --- | --- | --- | --- | --- |
| Age, years | -0.263 | 0.001 | -0.253 | 0.002 | 0.095 | 0.241 |
| Diabetes duration, years | -0.342 | <0.001 | -0.367 | <0.001 | 0.277 | <0.001 |
| Age at onset, years | 0.167 | 0.033 | 0.201 | 0.010 | -0.226 | 0.004 |
| Body mass index, kg/m<sup>2</sup> | -0.033 | 0.680 | 0.121 | 0.124 | -0.135 | 0.088 |
| Systolic blood pressure, mmHg | -0.028 | 0.738 | 0.005 | 0.954 | -0.084 | 0.322 |
| Diastolic blood pressure, mmHg | 0.158 | 0.044 | 0.156 | 0.047 | -0.172 | 0.029 |
| HbA<sub>1c</sub>, mmol/mol | -0.132 | 0.097 | -0.119 | 0.136 | 0.097 | 0.222 |
| Creatine, μmol/l | -0.228 | 0.005 | -0.176 | 0.031 | -0.084 | 0.306 |
| Total cholesterol, mmol/l | -0.006 | 0.935 | -0.005 | 0.947 | -0.032 | 0.683 |
| HDL cholesterol, mmol/l | -0.026 | 0.742 | 0.019 | 0.813 | 0.176 | 0.027 |
| LDL cholesterol, mmol/l | 0.055 | 0.481 | 0.006 | 0.939 | -0.110 | 0.166 |
| Triglycerides, mmol/l | 0.016 | 0.838 | 0.086 | 0.265 | -0.071 | 0.357 |
| eGFR, ml/min/1.73m<sup>2</sup> | 0.352 | <0.001 | 0.240 | 0.004 | -0.080 | 0.343 |
| hs-CRP, mg/l | -0.027 | 0.737 | 0.016 | 0.842 | -0.022 | 0.788 |
| CMBs, n | -0.141 | 0.065 | -0.160 | 0.037 | 0.045 | 0.557 |
| WMHs, n | -0.227 | 0.005 | -0.197 | 0.016 | 0.107 | 0.195 |

Univariate linear regression analyses showing standardized beta coefficients and corresponding p-values for the associations between various clinical, biochemical, and brain MRI parameters with superficial capillary plexus vessel density (SCP), deep capillary plexus vessel density (DCP), and foveal avascular zone (FAZ) among individuals with type 1 diabetes

CMB, cerebral microbleed; eGFR, estimated glomerular filtration rate; HDL, high-density lipoprotein; hs-CRP, high-sensitivity C-reactive protein; LDL, low-density lipoprotein; WMH, white matter hyperintensity.

### OCTA and cSVD markers among individuals with type 1 diabetes

SCP and DCP vessel densities were smaller among those with CMBs (45.1% vs 46.5%, p=0.034, and 49.9% vs 52.9%, p<0.001, respectively) (Figure 1). No difference in FAZ was observed between the groups (0.28 [0.21-0.37] mm^2^ vs 0.28 [0.22-0.37] mm^2^, p=0.816). Neither of the vessel densities nor FAZ differed between participants with and without WMHs. All results remained unchanged after excluding individuals with macular edema from the analysis (data not shown).

**Figure 1.**
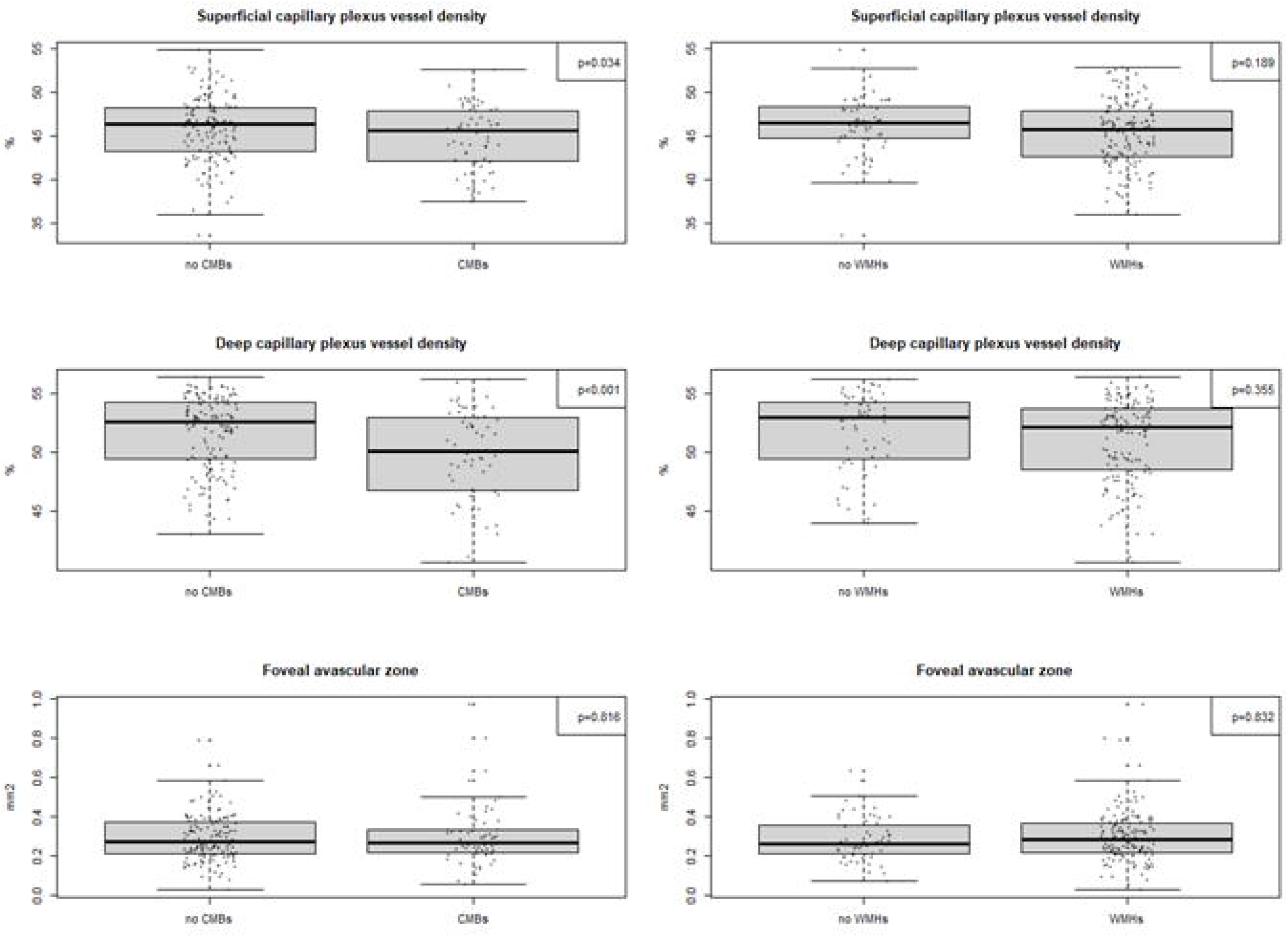
Boxplots showing group comparisons of retinal microvascular measures by the presence of cerebral microbleeds (CMBs) and white matter hyperintensities (WMHs). Superficial and deep capillary plexus vessel densities and foveal avascular zone areas are compared in individuals with type 1 diabetes.

**Figure 2.**
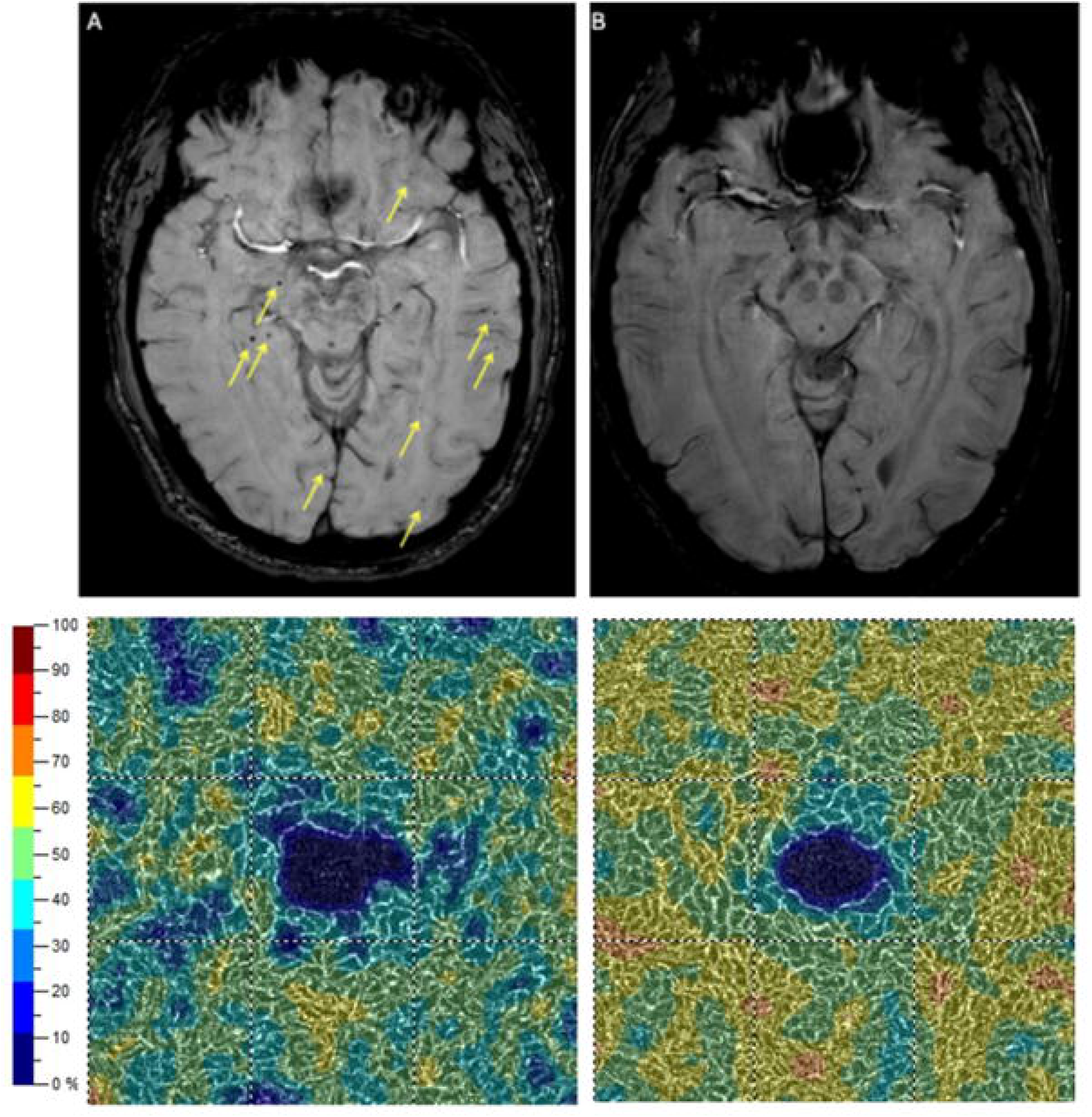
Example images from two individuals illustrating the association between cerebral microbleeds and retinal microvascular changes. Top row: susceptibility-weighted magnetic resonance imaging showing multiple cerebral microbleeds (yellow arrows) in individual A (left), and no detectable microbleeds in individual B (right). Bottom row: corresponding deep retinal capillary plexus vessel density maps obtained from optical coherence tomography angiography. Individual A, with numerous cerebral microbleeds, shows markedly reduced vessel density in the deep capillary plexus, particularly in the parafoveal region. In contrast, individual B, without cerebral microbleeds, demonstrates a more preserved and homogeneous vessel density pattern.

### Comparison between individuals with type 1 diabetes and healthy controls

No differences in SCP or DCP vessel densities or FAZ area were observed between individuals with type 1 diabetes and healthy controls (Table 1). DCP vessel density was slightly lower among those individuals with type 1 diabetes and CMBs vs healthy controls (p=0.031), but no difference in any other group comparison was observed in any other OCTA variable between healthy controls versus any cSVD, versus any CMB, or versus any WMH (Supplementary Figure 1).

As sensitivity analyses, we compared those who had a diabetes duration of at least 30 years with controls. Among those with longer diabetes duration, vessel densities of both SCP (44.4 [41.4-46.4] % vs 46.2 (44.0-47.0), p=0.006) and DCP (49.7 [47.0-52.9] %, p=0.001) were smaller, but no differences were found in FAZ area (p=0.052).

## Discussion

Our main finding was that neurologically asymptomatic individuals with type 1 diabetes with CMBs on brain MRI exhibited decreased vessel densities in both DCP and SCP, compared to those without CMBs. The FAZ area did not differ between these groups. Participants with and without WMHs showed no differences in retinal microvascular structure. In multivariate analysis, all three OCTA parameters correlated with the duration of type 1 diabetes; vessel densities negatively and FAZ positively, as expected. Finally, we observed no differences in OCTA parameters when comparing individuals with type 1 diabetes and healthy controls.

To the best of our knowledge, no previous study has investigated the associations between cSVD brain markers and microvascular retinal parameters in middle-aged individuals with type 1 diabetes. To find early vascular pathology, our sample included individuals without any signs or symptoms of neurological disease. Individuals with CMBs showed microvascular abnormalities in the retina compared to those without CMBs, manifested as lower SCP and DCP vessel densities. We observed no difference in FAZ area, which may be due to a larger variability in this parameter, compared to capillary plexus vessel densities, since FAZ is known to exhibit large variability even among healthy individuals ^22^. However, our findings align with a study conducted in older, on average 60-year-old individuals with type 2 diabetes, where decreased vessel density of the superficial vascular plexus was associated with CMBs ^5^. In the same study, when comparing those with and without cSVD, differences were found in DCP and superficial vascular plexus, but not in FAZ sizes ^5^. Correspondingly, contrary evidence exists, since some studies conducted in older individuals have found no associations between OCTA markers and CMBs ^13,16,17^. However, our current findings suggest that retinal microvascular changes might manifest an extension of vascular pathology in the brain.

There were no differences in OCTA markers in those with and without WMHs. These results differ from previous studies in older adults. Bermudez *et al.* showed associations between decreased retinal capillary densities in the superficial and deep fovea, as well as the superficial parafovea, and increased WMHs in individuals aged on average 75 years without dementia ^13^. Wang *et al.* found an association between WMHs and SCP vessel density in on average 64-year-old individuals diagnosed with cSVD ^14^. Another study found WMHs to be associated with decreased DCP vessel density in on average 60-year-old individuals with sporadic cSVD ^15^. One community-based study found WMHs to be associated with SCP, DCP, and FAZ in individuals with a median age of 68 years ^16^. One explanation for not finding associations between WMHs and OCTA parameters might be that the WMHs in our middle-aged participants were subtle (Fazekas 0 or 1). Enlarged FAZ has been shown to relate to higher Fazekas scores, supporting this hypothesis ^23^.

In the multivariate analysis, all three OCTA parameters correlated with the duration of type 1 diabetes but not with age or HbA_1c_. The treatment of diabetes has developed considerably in recent years, leading to improved glycemic control, which in turn has been speculated to reflect the smaller prevalence of diabetic retinopathy today ^24,25^. Thus, the cumulative pathological effect of diabetes-related factors on retinal microvasculature, such as longstanding hyperglycemia ^26^, has likely been greater than age, even though aging has also been shown to contribute to diabetic retinopathy ^27^. One explanation for not finding significant associations between HbA_1c_ values and OCTA markers could be that we used data from a single time-point, reflecting only the current glycemic status. We cannot, thus, rule out a potential role of long-term glycemic control values.

The finding that there were no differences in OCTA parameters between individuals with type 1 diabetes and healthy controls is unexpected but consistent with a meta-analysis by Zhang *et al.* ^28^. Overall, the evidence is contradictory. Earlier studies have reported differences in SCP but not DCP or FAZ ^29^, and DCP but not SCP or FAZ ^9^, between middle-aged individuals with type 1 diabetes and healthy controls. Some studies have identified microvascular retinal alterations with OCTA even in young adults with type 1 diabetes and no retinopathy ^30–32^. Similarly, decreased SCP and DCP vessel densities and enlarged FAZ have been reported in older individuals with type 1 diabetes without retinopathy ^10^. One factor contributing to these mixed findings may be the small sample sizes of many of the studies ^28,33^ and the use of different OCTA devices in different studies, since measured values appear to vary considerably between different scanners ^34,35^. Additionally, aging has been shown to affect retinal microvasculature also in healthy individuals and some studies have defined changes in capillary plexus vessel densities and FAZ to start manifesting after the age of 40 years ^36,37^.

The strengths of our study are the relatively large sample size, consistency of our methodology, strong phenotypes, and standardized and consistent MRI interpretations. However, certain limitations should be noted. One is the use of single HbA_1c_ measurements in the analysis; since the glycaemic control has improved in recent years, this may not reflect long-term glycaemic control so well. In addition, our control group had a surprising amount of cerebral small vessel disease for their age ^38,39^, which might influence our lack of findings in the comparison analysis type 1 diabetes vs controls. One reason for this might be that individuals with more subjective symptoms, e.g., cognitive complaints, may be more likely to volunteer for this type of study, leading to a sample with overrepresentation of cSVD.

## Conclusions

Although OCTA metrics did not differ between type 1 diabetes and controls overall, within the type 1 diabetes cohort, lower SCP and DCP vessel densities were associated with CMBs, and diabetes duration independently related to all three OCTA variables. These findings suggest that OCTA could serve as a pragmatic, non-invasive retinal surrogate for cerebral microangiopathy in long-duration type 1 diabetes, helping to prioritize patients for brain MRI and tighter vascular risk management, while cautioning against using OCTA to infer WMH burden.

## Declarations

### Ethics approval and consent to participate

The study follows the Declaration of Helsinki, with approval from the Ethics Committee of Helsinki and Uusimaa Hospital District (ID: HUS/2184/2017). Written informed consent was obtained from all participants.

### Consent for publication

All authors contributed to the interpretation of the results and gave their critical comments for the manuscript. All authors agreed on the final content

### Availability of data and materials

Individual-level data from the study participants are not publicly available due to consent restrictions provided by the participants at the time of data collection. Readers may propose collaboration to access the individual-level data by contacting the lead investigator.

### Authors’ contributions

AT, IK, MIE, PHG, LT, JAT, JM, JP, and DG contributed to the study design, acquisition and interpretation of data. JM interpreted MRI images and JAT OCTA images for macular oedema. AT prepared first draft of the study. All authors contributed to the interpretation of the results and gave their critical comments for the manuscript. All authors agreed on the final content. DG is the guarantor of this study and takes responsibility for the integrity of the data and the accuracy of the data analysis.

## List of abbrevations

CMB: Cerebral Microbleed
cSVD: Cerebral Small Vessel Disease
DR: Diabetic Retinopathy
DCP: Deep Capillary Plexus
FAZ: Foveal Avascular Zone
Finndiane: Finnish Diabetic Nephropathy Study
MRI: Magnetic Resonance Imaging
OCTA: Optical Coherence Tomography Angiography
SCP: Superficial Capillary Plexus
WMH: White Matter Hyperintensity

## Acknowledgements

We gratefully acknowledge the technical assistance of Anu Dufva, Anna Sandelin, and Kirsi Uljala. We also gratefully thank Pentti Pölönen, Department of Radiology, Helsinki University Hospital, for performing the MRI scans and Oili Salonen, Department of Radiology, Helsinki University Hospital, for her contribution in planning the MRI protocol.

## Figure legends

**Supplementary Figure 1.** Group comparison box plots between healthy controls and different subsets of individuals with type 1 diabetes. T1D = Type 1 diabetes, CSVD = cerebral small vessel disease, CMB = cerebral microbleed, WMH = white matter hyperintensity

