## Supplementary figures and images for "Relations between retinal microvasculature by optical coherence tomography angiography and cerebral small vessel disease in individuals with type 1 diabetes"

### Supplementary figure 1

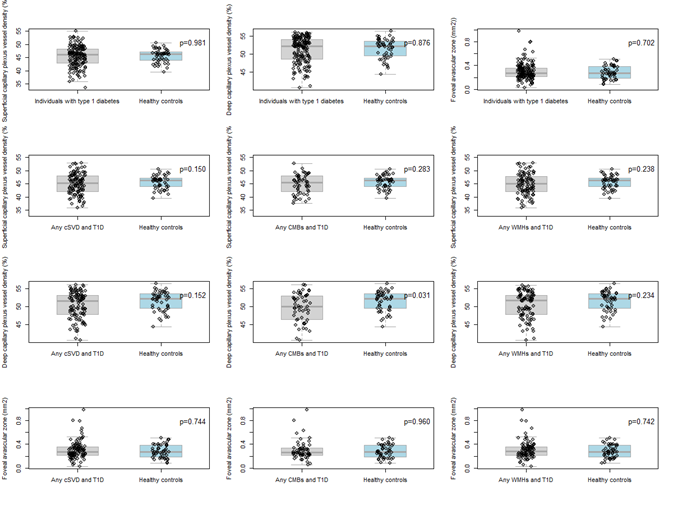
